# Digitally-enabled self-experiments in general practice for evaluating the effectiveness of non-pharmacological, self-management interventions for persistent pain: Protocol for single case experiments

**DOI:** 10.1101/2024.12.20.24319270

**Authors:** Nancy Sturman, Jane Nikles, Peter Worthy, Shauna Fjaagesund, Rachel Elphinston, Nicole Andrews, Michele Sterling, Stefan Konigorski

## Abstract

**Background:** Non-pharmacological, self-management interventions are often recommended for general practice patients with persistent pain, a distressing, costly and heterogenous condition with variable responses to treatment interventions. Single-case experimental designs (“self-experiments”) evaluate intervention effectiveness at the individual level. StudyU is an open-source digital platform designed to help patients undertake these self-experiments.

**Objectives:** To investigate the feasibility and acceptability (for patients, practitioners and other practice staff) of integrating digitally-enabled, withdrawal/reversal self-experiments in the general practice care of patients with persistent pain.

**Methods:** We will recruit 50 patients from a large Australian general practice. Participants will trial a self-selected, self-management intervention (such as a physical activity, mindfulness practice, or online, self-guided cognitive behavioural therapy) approved by their general practitioner (GP), and use the StudyU app to rate the daily impact of their pain over the 10-week study period. The primary clinical outcome of the self-experiments is pain interference (measured using the modified Brief Pain Inventory) and tested for its mean difference between usual routine and intervention conditions. Clinical reports are generated for the patient and their GP.

We will use validated measures of app usability and acceptance, pre/post measures of patient self-efficacy, quality of life, health service use and self-reported health, individual interviews informed by Normalisation Process Theory, and a nested process evaluation to examine the feasibility and acceptability for patients and practice staff of embedding these self-experiments in general practice care.

**Results:** The research was funded in September 2023, funding and other agreements were completed by December 2024, and patient recruitment is scheduled to start in January 2025.

**Conclusion:** Digitally-enabled self-experiments testing non-pharmacological treatment effectiveness may empower patients to self-manage persistent pain and adopt personally effective non-pharmacological interventions, in partnership with their GPs, and provide a model for integrating other new technology into the general practice care of patients with other chronic conditions.

## INTRODUCTION

### Persistent pain

Innovative and practical solutions are needed to address the costly and distressing problem of persistent (or chronic) non-cancer pain. Living with persistent pain is associated with substantial disability (1) and healthcare system cost (2). Persistent pain also presents challenges for general practice, including navigating ongoing compensation and insurance matters, and patient dependence on pharmacological interventions (3). Although non-pharmacological, self-management and resilience-building options for managing pain are often recommended, persistent pain is a highly heterogenous condition with variable individual responses to treatments (4). Single-case experimental designs (personalised, single patient self-experiments using patients as their own controls) (5) are well positioned to evaluate effectiveness at the individual level (4), but there is very limited evidence about the feasibility and acceptability of adopting these in general practice.

### Digitally enabled self-experiments: StudyU

The open access StudyU app is a mobile phone app designed for conducting Single Case Experimental Designs (6). In this study we use the app to enable patients with persistent pain to evaluate the effectiveness of their choice of a self-management, non-pharmacological intervention. We have piloted the app with four community-based consumers with persistent pain. The app is embedded in the StudyU platform (6), which also includes a general-use tool for researchers and healthcare practitioners to design, monitor and manage these patient self-experiments.

The StudyU app will generate daily reminders for patients to undertake their chosen activity, and rate pain severity and impact, over a 10-week test period. After patients complete their self-experiments, the app will provide visual displays of daily ratings, and average baseline versus intervention ratings, using colour-coded bar graphs. The research team will also analyse outcomes using t-tests and Bayesian linear mixed models to compare baseline and intervention conditions, producing clinical reports which are forwarded by a secure messaging system to the patient and their GP. StudyU does not require a user account and does not store any personal identifying data. The anonymized recorded study data will be published after study completion to contribute to an open worldwide data repository, with patient consent, to allow aggregation of single case experiments testing similar interventions in future open science approaches, and inform the design of future trials.

### Apps in general practice

Apps are used in general practice to improve consumer adherence (7), monitor symptoms (8), supplement medical histories (9), and implement management algorithms (10). Over 50% of Australian general practitioners (GPs) in a 2019 survey study recommended mHealth apps to patients at least monthly (11). However, GPs also perceive barriers to effectively adopting evidence-based apps, including limited awareness of suitable apps, and concerns about time commitment, privacy, safety, and trustworthiness (12). Australian patients appear less concerned than GPs about privacy and data safety issues, and appreciate their doctors recommending evidence-based apps (12), although social and economic disadvantage (particularly low income, education and employment) and rural location may reduce digital access, affordability and ability (13, 14). In international literature, workflow adjustments, inadequate reimbursement and high training effort are substantial barriers for digital health adoption by GPs, whereas interoperability, integration with workflow, continued technical support, improved usability, digital formularies, payment models and attention to personal and emotional elements facilitate uptake (15, 16). The engagement of both health professionals and patients is essential for successful integration (12).

#### Aim

The overall aim of this research is to empower patients with persistent pain and their treating teams to adopt effective self-management activities and discontinue treatments which are ineffective for them personally (even if these are generally recommended).

#### Research question

Is it feasible, acceptable and useful to embed digitally-enabled self-experiments into a general practice setting, to test the effectiveness of non-pharmacological treatments in patients with persistent non-cancer pain?

## METHOD

### Design

#### Setting and Participants

The study is conducted in a large privately-owned general practice north of Brisbane, Australia. The practice serves a predominantly low socio-economic patient demographic, and has a commitment to practice-based research that improves patient care in their community. There were 5,516 presentations for pain as the primary reason for attendance in 2020-2021 at the practice. The practice team has worked with us to design operational procedures which embed StudyU-enabled self-experiments in chronic disease and pain management consultation workflows and systems. We will recruit four GPs, four practice nurses and up to 50 patients with a diagnosis of persistent pain, and estimate that 30 patients will complete the study, an acceptable number for a feasibility study (17).

##### Inclusion criteria

1) >18 years, 2) currently experiencing clinically significant persistent pain for 3 months or longer, most days per week (average pain severity in last week of at least 3-4/10), 3) on stable dose/s of regular pain medication (including medicinal cannabis) for >/= 4 weeks prior, or not currently taking pain medication.

A sub-group of participants will be recruited with an additional inclusion criterion of “persistent neck and/or back pain following a road traffic crash at least 3 months prior to recruitment”.

##### Exclusion criteria

1) acute mental health disorder or suicidal, 2) unable to use digital health apps due to impairments in cognition, vision or dexterity 3) non-English speaking, 4) no access to smartphone or Internet, 5) recent (in last 4 weeks) or planned (in the next 3 months) changes to current pain management interventions, including surgery.

#### Research Plan

A patient flyer about the study, and a one-page explanatory information sheet, will be displayed in practice waiting areas, and consulting rooms. Study GPs and/or practice nurses will provide further information to patients who express an interest in participating, screen them for study eligibility, invite eligible patients to provide written, informed consent to participate, assist patients to select a suitable non-pharmacological intervention, and complete study referral information. See Figure 1 for an overview of participant flow and study design. A steering committee will oversee and advise the operational research team (the Project Manager (JN), the research assistant (SF) and the Principal Investigator (NS)).

**Figure 1.**
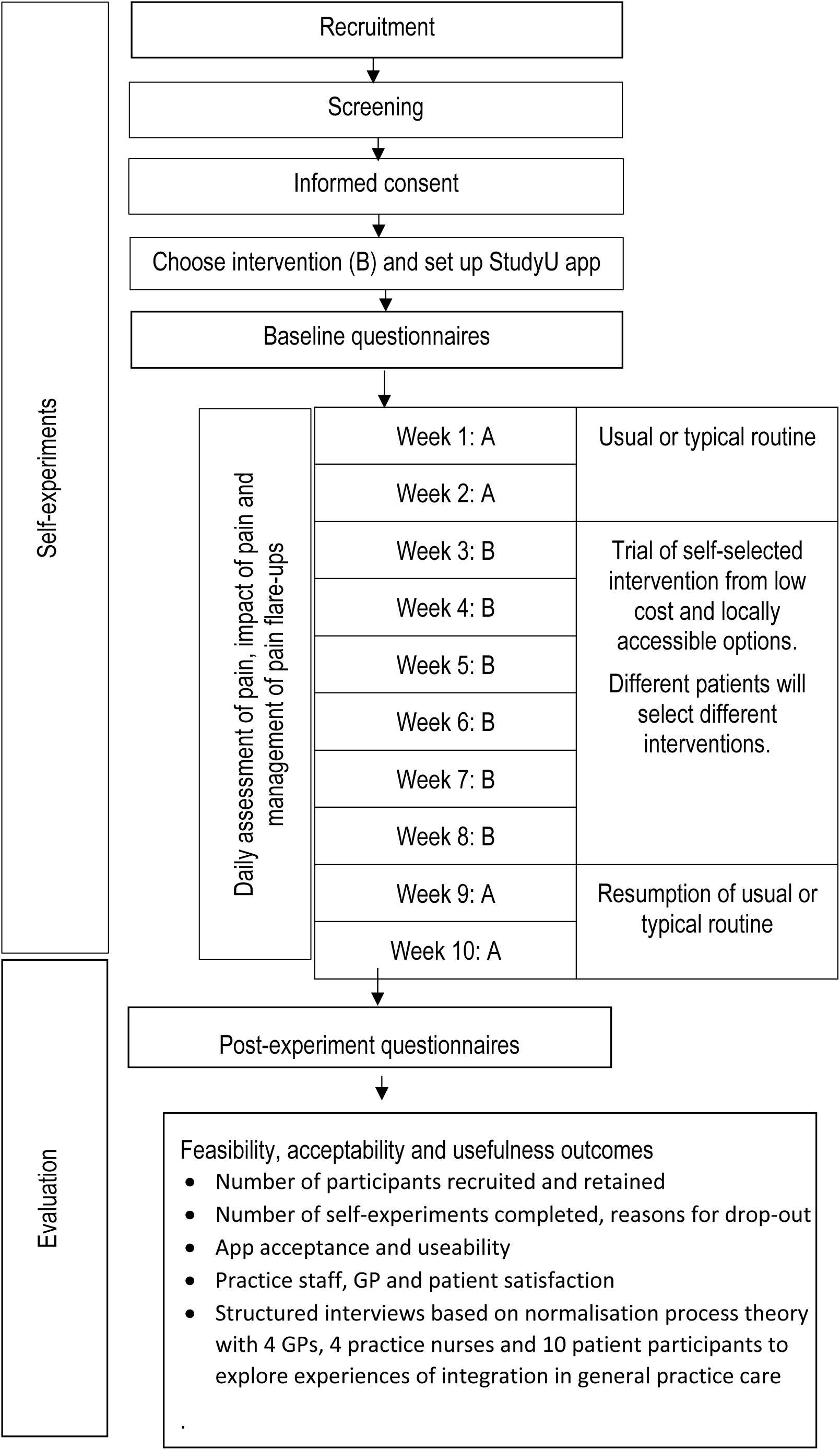
Overview of participant flow and study design

#### Intervention

A research assistant embedded within the participating general practice will prepare, and regularly update, a list of low-cost activities which are generally recommended for the management of persistent pain and are locally available at low or no cost to practice patients. The activities are reviewed by a pain clinician to ensure they are generally appropriate. The list includes mindfulness practice, online pain management modules, low impact exercise such as chair yoga or tai chi, walking programs (self-managed or group), and personalised or small group exercise programs. The patient selects an activity from this list, with the approval of their GP. On receipt of the study referral, which is generated and auto-populated from existing practice software, the research assistant designs a personalised self-experiment for each patient using the StudyU designer, and forwards an invitation code to the patient, for them to enter into the StudyU app to commence their personalised self-experiment.

Patients are assisted by the study team or practice nurses to download the StudyU app and enter their personalised invitation code. The patient’s self-experiment will continue for a total of 10 weeks, using an A^1-^B-A^2^ withdrawal/reversal single case experiment design, where Phase A^1^ is the patient’s usual or typical routine, Phase B is 6 weeks undertaking the intervention activity, and Phase A^2^ is resumption of usual routine (except for patients who wish to continue the intervention activity).

After the patient has completed their study, they will be encouraged to book a consultation with their GP to discuss study results. This consultation will include shared decision-making about whether or not to continue the activity. Patients may also choose to conduct a second self-experiment with another activity.

Gift vouchers will be provided to practice nurses to reimburse them for time spent recruiting and assisting patients. Study nurses and GPs will also be reimbursed for attendance at a training session to familiarise them with study rationale and procedures, and participating in individual interviews at study completion. Gift vouchers will be provided to patients to reimburse them for the initial study visit, survey completion and post-study interviews.

#### Outcome measures

The primary clinical outcome measured daily in the app-enabled self-experiments is pain interference, using the modified Brief Pain Inventory 7-item interference subscale (18). A change of 1 point over the average of the 7 items is defined as clinically significant (19). The secondary clinical outcomes are pain intensity, using a Visual Analog Scale (20), and number of additional treatments for pain flare-ups. All clinical outcomes are measured in the StudyU app.

In addition to these clinical outcomes, patients will also complete validated measures at baseline, and 4 weeks following their self-experiment, of self-efficacy, mental health, self-reported health status, health service use, and quality of life (see Box 1 for full list of instruments used).

We will examine the feasibility and acceptability of integrating digitally-enabled self-experiments over the 12-18 months’ study duration, using a nested process evaluation, and structured individual interviews, informed by Normalisation Process Theory Murray (21), with study GPs and practice nurses following study completion. Patients will complete measures of technology acceptance and usability (see Box 1) and participate in structured interviews after their self-experiments, exploring their experiences. Feasibility outcome measures include number of participants recruited and retained, and self-experiment completion rate. All management decisions and reasons for drop-out will be recorded by the research team. Outcome measures are summarised in Box 1.

#### Data Storage and Data Privacy

As described above, different types of data will be assessed during the study. Questionnaire data from baseline and after 4 weeks as well as Interview data will be collected and stored locally at University of Brisbane. All data from the single-case experimental designs will be recorded using the StudyU app. As no user accounts are needed for using the StudyU app, no identifying data is collected through the StudyU app. In order to link the data from the single case experimental designs to further data collected in baseline questionnaires, participants enter the StudyU app using an invite code which acts as a pseudonym. All data collected through the StudyU platform is stored on secure servers at the Hasso Plattner Institute in Germany, and will be published openly through the StudyU Designer website (https://designer.studyu.health) after study completion and after deletion of the invite codes to guarantee anonymized data. This will contribute to an open worldwide data repository, allow aggregation of single case experiments testing similar interventions in future open science approaches, and inform the design of future trials.

#### Statistical Analyses

For study planning and for statistical analyses, aims for individual-level and population-level analyses can be distinguished.

In order to answer primary research questions regarding feasibility and usability, we will analyse data collected across participants and provide descriptive statistics that summarise usability, usefulness, acceptability and feasibility outcomes. In order to assess the effectiveness of clinical interventions on the individual level, we aim to test whether the primary and secondary clinical outcomes would differ between phases when the intervention was applied and not applied (standard routine), for each individual separately. For this, we will perform t-tests comparing average daily scores of primary and secondary clinical outcomes between usual routine and intervention phases, as well as Bayesian linear mixed models to calculate the posterior probability that the intervention is effective at the individual level. We will consider two definitions of treatment responders: having an estimated posterior probability higher than 80% of a reduction by at least 1 unit in pain interference (i.e., in the average of the 7 items in the modified Brief Pain Inventory 7-item subscale (19)), and of any reduction in pain interference, respectively. It may be possible to perform these analyses at an aggregate level across patients, if multiple participants test the same intervention. In all analyses, the primary analysis will assume data is missing completely at random and perform analyses of the complete-case data (for each individual, separately), without imputation. Sensitivity analyses will test the robustness of the results after multiple imputation of missing data. For the clinical reports, we will report the results both from t-tests and Bayesian linear mixed models. All analyses will be performed using the statistical software R. Structured interviews will be transcribed and analysed thematically using template analysis (22).

As our primary aim is to evaluate feasibility of embedding self-experiments in general practice, we did not perform statistical sample size calculations for study planning. Also regarding self-experiments, our primary aims are in testing feasibility of reporting the results to clinicians and patients. Regarding the reported results of the trials, the effectiveness of interventions will be tested at the individual level. For these analyses, 70 measurements (one daily measurements over 10 weeks) will be available for each patient if there is no missing data. For a patient with 20% missing data, a two-sided two-sample t-test at significance level 0.05 would have a power of identifying treatment effects that have an effect size of Cohen’s D of at least 0.53 with a power of 80%.

## RESULTS

The research was funded in September 2023, by the MBHF-RACGP Foundation 2023 and MAIC-RACGP Foundation 2023 grants. Full funding and other agreements were completed by December 2024. Practice protocols have been developed, and practice staff have received training in the StudyU app and patient recruitment. Patient recruitment is scheduled to start in January 2025. Results are expected by December 2025. See Figure 2 for screenshots of the StudyU app, and Figure 3 for the project Logic Model, including inputs, proximal outcomes and distal outcomes.

**Figure 2.**
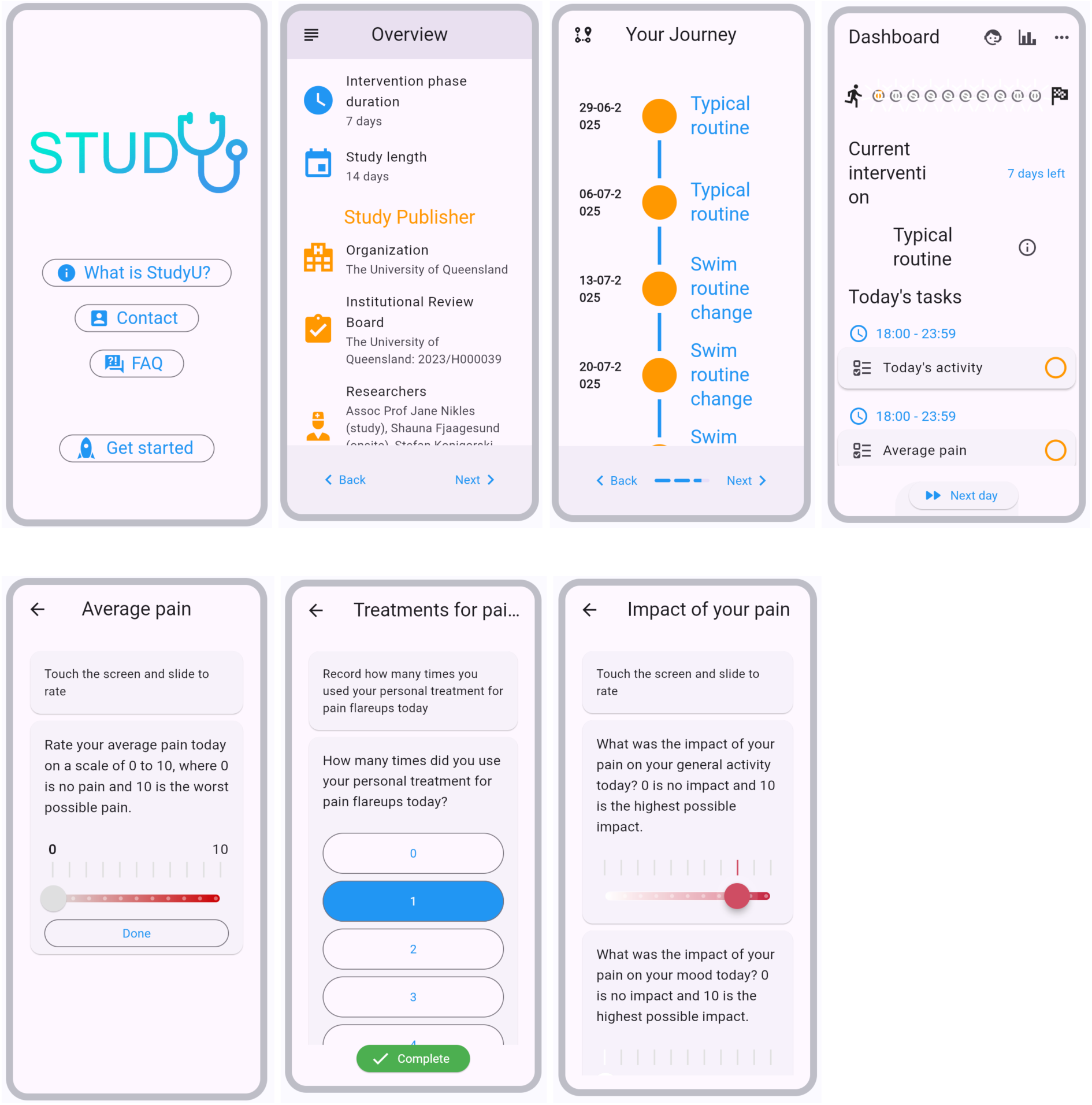
Illustrative screenshots

**Figure 3.**
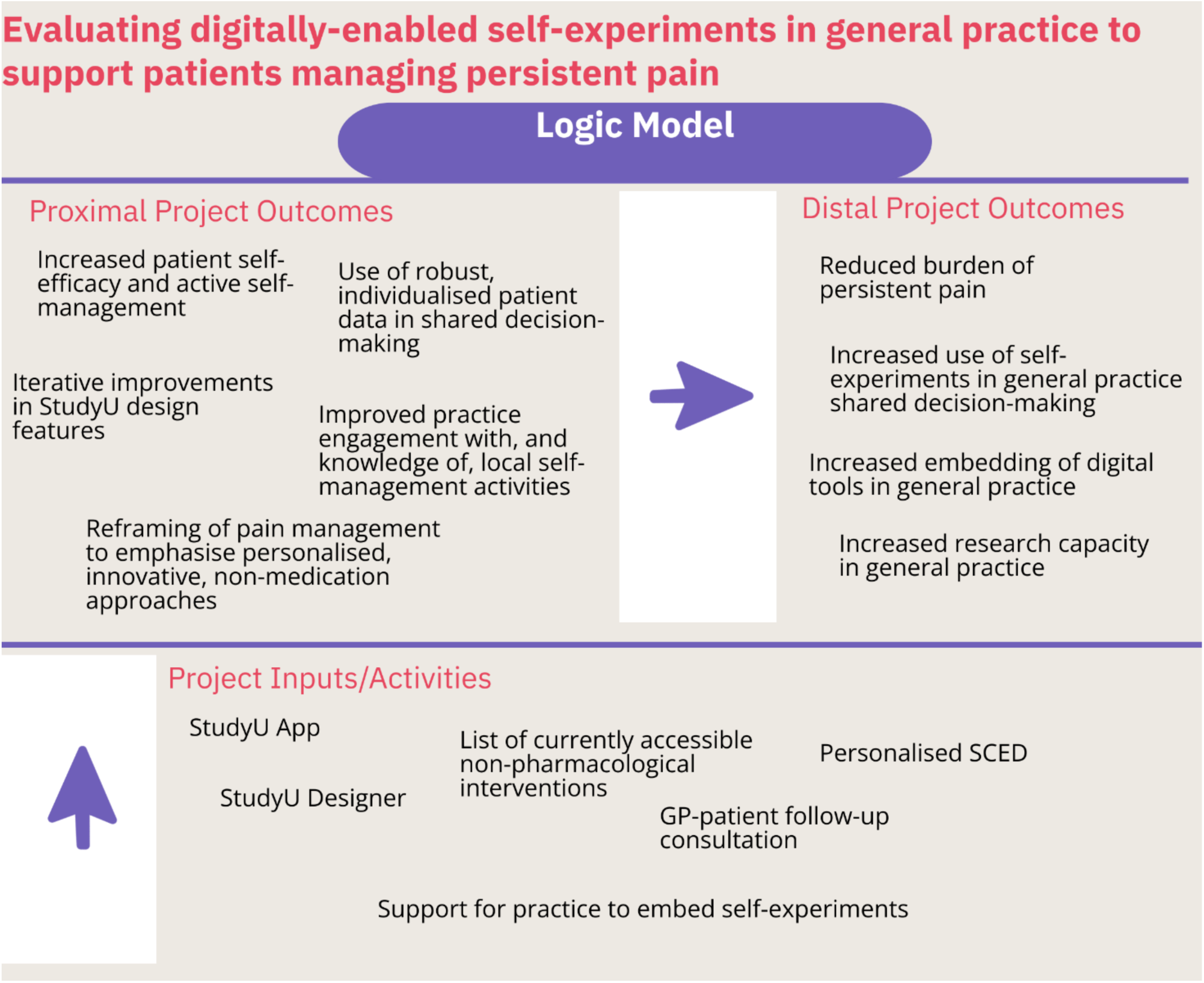
Project Logic Model

## DISCUSSION

### Findings

In addition to the clinical benefits for individual patients of identifying effective (or ineffective) treatments, study findings will cast light on the feasibility, acceptability and usefulness of embedding digitally-enabled single case experiments as a new tool for GPs to support patients with persistent pain.

### Strengths

Single case experimental designs, despite some limitations, provide more robust evidence of intervention effectiveness than the more common “try it and see” approaches in general practice (23). Digitally-enabled single case experiments are a novel approach in general practice. Patients are able to choose an intervention which is personally attractive, and we anticipate that use of the StudyU app will facilitate adherence to the planned intervention through the daily prompts and patient interest in establishing its effectiveness, potentially enhancing patient self-efficacy in relation to the self-management of their pain. We have addressed several known barriers to integration into general practice care, by embedding the self-experiments in existing practice workflows and systems, and facilitating patient referrals to locally accessible and affordable community-based services and activities

### Limitations

The relatively short duration of the self-experiments limits our ability to detect any intervention effect with a delay in onset longer than 3-4 weeks, or assess longer term effects (beyond 4 weeks after experiment completion) of the intervention. The ABA withdrawal/reversal, single baseline design does not include within-case replication, assessment of baseline stability, blinding, randomisation and does not include additional blocks that may prevent wash-in (delayed onset) or wash-out (carry-over) effects of the interventions. We considered incorporating some of these design features, in order to increase the validity of self-experiment results (24), but considered that adding further to the length and complexity of the self-experiments was likely to reduce patient interest and engagement in our pilot feasibility trial. However, we exclude patients with recent or intercurrent changes in other pain management interventions, or with cancer pain (which may be progressive). Limitations and caveats can be discussed during the follow-up GP-patient consultation, and future studies could optimise the length and design of trials for particular interventions. We are also testing StudyU-enabled self-experiments in a relatively small study in a single practice that is already research-friendly and has provided funds to support the initial pilot: this limits the generalisability of our feasibility results to other practices which may have additional barriers to successfully integrating StudyU into clinical care. Patients self-select into the study, non-English speakers are excluded, and we have not provided devices, data or internet access as strategies to enhance engagement, reducing patient diversity and representativeness. The restrictions are important for generalizations of the study results, in particular, it would not possible to conclude that digitally-enabled self-experiments are appropriate for all patients with persistent pain. The self-selection of participants interested in performing the trials is rather a feature than a limitation, as we believe that studying the effectiveness in this particular group is of interest. We have not attempted to standardise or fidelity-check selected interventions or self-reported behaviours, which may make it more difficult to aggregate these single case experiments in future studies.

### Implications for practice

In addition to the implications for patients with persistent pain, findings may also contribute to a model for integrating other technology and single case experimental designs into the general practice care of patients with other chronic conditions.

## Data Availability

All trial data collected in the study will be made publicly available at https://studyu.health after completion of the study.

## Funding

Royal Australian College of General Practitioners Foundation, Medibank Better Health Foundation and Motor Accident Insurance Commission Qld have generously funded this study. MS receives a fellowship from the NHMRC (APP2017405) and receives unrestricted funding from the Motor Accident Insurance Commission of Qld.

## Ethical approval

The University of Queensland Human Research Ethics Committee 2023/HE000039.

## Australia and New Zealand Clinical Trials Registry

ACTRN12624000459527

## Competing interests

Jane Nikles has a commercial interest in N-of-1 Hub Pty Ltd consultancy company.

## Acknowledgements

We acknowledge Health Hub Doctors Morayfield, in particular Assoc Prof Evan Jones, Dr Shahab Sojoudi Haghighi, and Dr David Shahar, for their support and enthusiasm for this research. We gratefully acknowledge our funders, the Royal Australian College of General Practitioners Foundation, Medibank Better Health Foundation and the Motor Accident Insurance Commission. We acknowledge Dr Suzanne McDonald for advice and input.

## Boxes

**Box 1.**
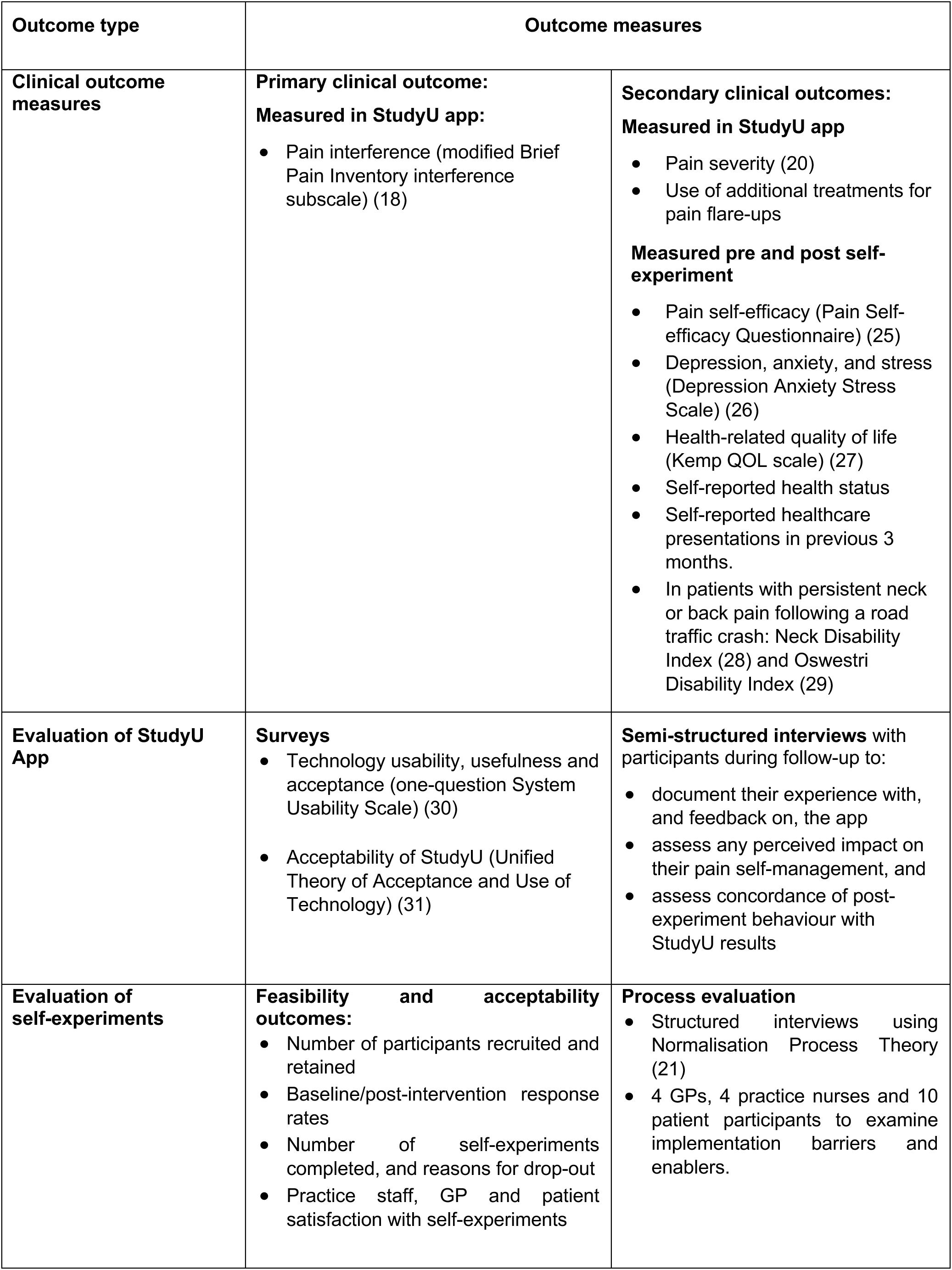
Summary of Outcome Measures.

## Notes

### Clinical Trial

Australia and New Zealand Clinical Trials Registry: ACTRN12624000459527

### Author Declarations

Ethical approval: The University of Queensland Human Research Ethics Committee 2023/HE000039.

### Summary of Updates

Manuscript has been updated, and more details have been added regarding the study protocol.

